# Who Falls After Stroke? Evidence From a Prospective Stroke Cohort

**DOI:** 10.64898/2026.01.08.26343741

**Authors:** Anna Kufner, Yunyou Tang, Uchralt Temuulen, Abbas Ghadir, Torsten Rackoll, Ulrike Grittner, Daniel Kroneberg, Benedikt Weigel, Andrea A Kühn, Martin Reich, Alexander H Nave, Matthias Endres

**Affiliations:** Charité – Universitätsmedizin Berlin, corporate member of Freie Universität Berlin and Humboldt-Universität zu Berlin, Klinik für Neurologie mit Experimenteller Neurologie, 10117 Berlin, Germany; Charité - Universitätsmedizin Berlin, corporate member of Freie Universität Berlin and Humboldt-Universität zu Berlin, Center for Stroke Research Berlin (CSB), 12203 Berlin, Germany; Charité – Universitätsmedizin Berlin, corporate member of Freie Universität Berlin and Humboldt-Universität zu Berlin, Institut für Biometrie und klinische Epidemiologie, 10117 Berlin, Germany; Berlin Institute of Health at Charité – Universitätsmedizin Berlin, 10117 Berlin, Germany; German Center for Cardiovascular Research (Deutsches Zentrum für Herz Kreislauferkrankungen, DZHK), Partner Site Berlin, 10117 Berlin, Germany; German Center for Mental Health (Deutsches Zentrum für Psychische Gesundheit, DZPG), Partner Site Berlin, 10117 Berlin, Germany; German Center for Neurodegenerative Diseases (Deutsches Zentrum für Neurodegenerative Erkrankungen, DZNE), Partner Site Berlin, 10117 Berlin, Germany; Berlin Institute of Health at Charité – Universitätsmedizin Berlin, QUEST Center for Responsible Research, 10178 Berlin, Germany; Department of Neurology, University Hospital and Julius-Maximilians-University, Wuerzburg, Germany

**Keywords:** Stroke, Falls, Mobility, Gait, Lesion network mapping

## Abstract

**Background:** Falls are a frequent and serious complication after stroke, affecting more than 30% of survivors within the first year. While age and comorbidities are established risk factors for falls, stroke-specific contributors—particularly lesion-related impairments in mobility and gait—are less well understood and may inform targeted secondary prevention.

**Methods:** We analyzed data from 94 patients with disabling subacute ischemic stroke enrolled in the prospective BAPTISe cohort, a predefined imaging and biomarker sub-cohort of the multicenter PHYS-STROKE trial. Detailed gait and mobility assessments were performed at baseline. Principal component (PC) analysis reduced seven mobility-related and four gait-related variables into two composite scores: PC1-Mobility and PC1-Gait, explaining 56% and 82% of variance, respectively. PC1-Mobility reflected global disability and functional mobility in daily life, whereas PC1-Gait captured spatiotemporal walking capacity and efficiency. Lesion network mapping (LNM) using a normative connectome identified functional networks associated with each domain. Patient-reported falls up to six months post-enrollment were the primary endpoint.

**Results:** LNM of PC1-Mobility revealed a predominantly cortical network involving pre- and postcentral gyri, superior and middle frontal gyri, and sensorimotor integration areas. In contrast, PC1-Gait was associated with subcortical and infratentorial connectivity, including bilateral thalamus, brainstem, and cerebellum. In multivariable regression, network similarity scores were not independently associated with falls; only older age was significant (adjusted OR1.08, 95%CI1.02–1.15,p=0.013). LNM of fall occurrence showed a cortical network with significant spatial overlap with the PC1-Mobility network(p<0.001).

**Conclusion:** This exploratory, hypothesis-generating study identified distinct lesion-derived functional networks associated with post-stroke mobility and gait impairment. Our findings suggest that falls may be more closely linked to disruptions in cortical networks involved in voluntary motor control and whole-body coordination, rather than subcortical structures primarily modulating gait. These results provide a foundation for future research aimed at improving fall risk stratification and targeted prevention strategies in stroke survivors.

## Introduction

Over one-third of stroke survivors experience at least one fall within the first year after stroke, with approximately one-third of these falls requiring medical intervention^1^. Falls in this patient population are associated with prolonged hospitalization, increased morbidity and mortality, and a significant decline in post-stroke quality of life^1,2^. Despite their clinical relevance, however, falls in stroke survivors specifically remain understudied with no evidence of effective therapy for secondary prevention of falls after stroke^3,4^.

Previous observational studies have identified several risk factors for falls in stroke populations, including advanced age, sedative medication, neuropsychiatric symptoms such as depression and cognitive impairment, and the degree of functional disability^1,5,6^. Reported risk factors in stroke survivors largely overlap with those observed in an age-matched general population^3^. However, certain stroke-specific factors—such as unilateral weakness, impaired coordination, and gait dysfunction—are likely to play a disproportionately larger role in post-stroke falls^7^.

Ischemic stroke can disrupt critical motor pathways, leading to both general mobility impairment and more nuanced gait dysfunction in up to 50% of stroke survivors^8^. Despite its clinical relevance, predicting individual fall risk remains difficult. Previous lesion studies suggest that damage to the corticospinal tract and adjacent structures in the capsular-putaminal region plays a role in supraspinal gait control^9,10^. Similarly, a recent lesion-network mapping study identified a so-called ‘gait network’, in which lesions functionally connected to the anterior cingulate cortex, midbrain, and pons were associated with gait impairment^11^. Given the involvement of multiple systems beyond the corticospinal tract, and substantial interindividual differences in compensatory capacity, lesion location alone is likely insufficient to fully explain or predict fall risk^10–12^. Collectively, our understanding of the neuroanatomical basis and complexity of post-stroke motor and gait dysfunction remains limited—particularly regarding their specific contribution to fall risk. A better understanding of stroke-specific risk factors for falls could improve identification of high-risk fallers and lead to targeted prevention strategies and individualized rehabilitation planning.

Therefore, we here analyzed a well-characterized prospective cohort of patients with disabling ischemic stroke who underwent mobility and gait performance assessments at study enrollment and were monitored for fall occurrence over a six-month period. The mobility domain captured global disability and basic functional mobility in daily life, whereas the gait domain represented spatiotemporal aspects of walking capacity and efficiency. Our objectives were to: (1) apply lesion-network mapping to identify networks associated with impaired mobility, gait, and falls; and (2) determine whether lesion-induced disruption to these networks is associated with the risk of falling after stroke.

## Materials and methods

### Participants

This study is an exploratory, secondary analysis of data from the prospective observational cohort BAPTISe (Biomarkers and Perfusion-Training-Induced Changes After Stroke; ClinicalTrials.gov identifier: NCT01954797^13^) embedded in the multicenter, randomized controlled trial PHYS-STROKE (Physical Fitness Training in Patients with Subacute Stroke; ClinicalTrials.gov NCT01953549^14^). Patients were enrolled if they had experienced a subacute ischemic stroke (5–45 days after symptom onset) and had a Barthel Index score ≤65 at screening, with the primary aim of evaluating the effects of aerobic exercise on mobility and activities of daily living during the subacute post-stroke phase. Patients underwent clinical assessments at study enrollment (baseline), directly after the 4-week intervention period, and at six-month follow-up. For inclusion in the present analysis, participants were required to have available pre-intervention magnetic resonance imaging (MRI). All participants provided written informed consent. The study was conducted by the Declaration of Helsinki and approved by the local ethics committee in Berlin (EA1/137/13). This study follows the STROBE guidelines for the reporting of observational studies^15^.

### Imaging

All patients received a standardized stroke imaging protocol with a 3T MRI scanner, as detailed in the original study protocol^13^. Lesions were manually delineated on fluid-attenuated inversion recovery (FLAIR) images by a neurologist using the semi-automated lesion masking toolbox Clusterize^16^. All lesion masks were reviewed by an expert radiologist. The resulting lesion masks were co-registered to brain-extracted and bias-corrected b0 images using the Brain Extraction Tool (BET) from the FMRIB Software Library (FSL)^17^. Lesion masks were then normalized to MNI152 standard space (1×1×1 mm resolution)^18^ using the Symmetric Normalization (SyN) algorithm implemented in the Advanced Normalization Tools (ANTs) for Python^19^.

### Clinical assessment

Our primary clinical endpoint was the occurrence of patient-reported falls during the six-month period following study enrollment. Falls were assessed during scheduled follow-up visits, where treating physicians asked participants whether they had experienced a fall since the last contact. While falls could occur during the 4-week intervention period, no falls were reported during supervised intervention sessions. Outside of these visits, fall ascertainment was not systematically recorded. Fall occurrence was recorded as a binary variable. To investigate potential neural correlates of fall risk, we focused on two key baseline domains: mobility and gait, which served as behavioral inputs for subsequent imaging analyses.

Mobility was assessed using a composite of seven clinical variables that capture overall functional status and general mobility. These included the Modified Rankin Scale (mRS), a clinician-rated scale ranging from 0 (no symptoms) to 6 (death) in which functional dependence is assessed; the EQ-5D-5L mobility subdomain, a patient-reported scale from 1 (no problems) to 5 (unable to walk); and the Rivermead Mobility Index (RMI), which evaluates the ability to perform functional tasks such as transfers, walking, and stair climbing (range: 0–15). In addition, we recorded scores on three mobility-related items from the Barthel Index—walking, stair climbing, and transfers—with higher scores indicating greater independence. Finally, use of a walking aid (yes/no) during walking test was documented.

Gait was assessed through four performance-based parameters, reflecting both short-distance and endurance walking ability. The 10-Meter Walk Test (10MWT) captured the number of steps, the time to complete the test, and calculated gait speed (meters/second). The Six-Minute Walk Test (6MWT) measured the total distance walked in six minutes, reflecting submaximal aerobic capacity and walking endurance.

### Data Preprocessing and Dimensionality Reduction

For a flow-chart depicting our analysis pipeline from data pre-processing to imaging analyses, refer to **Figure 1**. All preprocessing and dimensionality-reduction steps with principal component analysis (PCA) were performed separately for the mobility domain (seven variables: mRS, RMI, EQ-5D-5L mobility sub-domain, and four Barthel sub-scores) and the gait domain (four variables: 10-m walk speed, number of steps, walking time, and 6-min walk distance).

**Figure 1:**
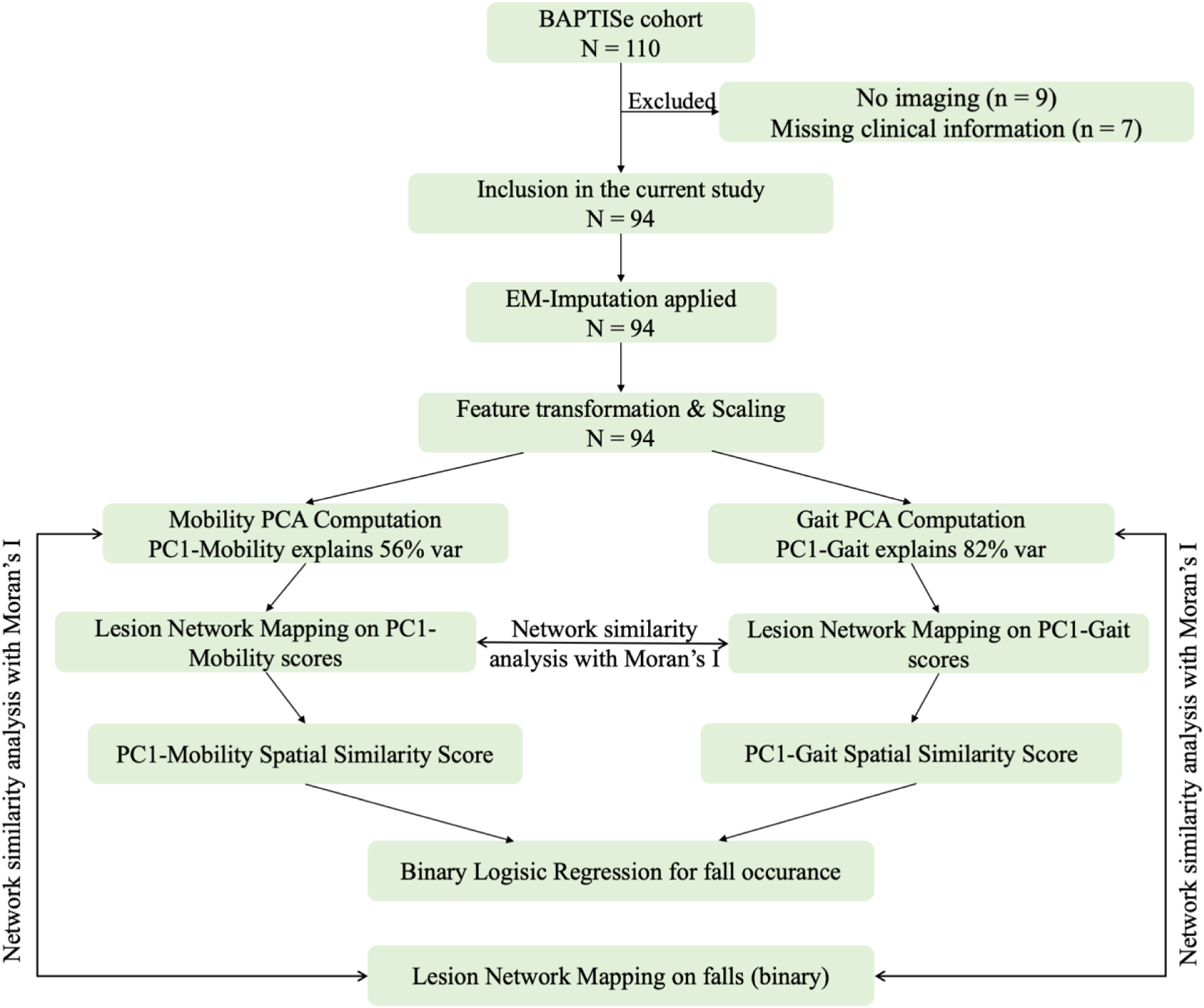
Flow-chart of the data analysis pipeline.

To harmonize directionality of the clinical variables, each variable was transformed so that higher values reflected worse function and zero corresponded to clinically normal performance. Accepted clinical reference values (e.g., 300 m for the 6-min walk, 1.2 m s⁻¹ for gait speed^20,21^) or full-scale maxima for ordinal measures were subtracted; negative residuals—representing better-than-normal performance—were truncated to zero to avoid artificial inverse correlations. For a detailed list of how all mobility and gait variables were transformed, refer to **Supplementary Table S1.** All variables were then z-normalized.

Because all variables were rescaled this way, we expected the first principal component (PC1) of each domain to correlate positively with every original metric. To test this assumption, we calculated Pearson correlations between PC1 scores and each variable and visualized the relationships (**Supplementary Figure S1** for mobility and **Supplementary Figure S2** for gait). Missingness across the 11 gait- and mobility-related variables ranged from 0% to 14.9 % (median ≈ 1%); the 6-Minute Walk Test distance had the most missing entries (n = 14). Missing entries were imputed via median-initialized expectation–maximization (EM; 50 iterations, tol = 1 × 10⁻⁴) implemented in Python 3.11 (scikit-learn 1.4), an approach shown to preserve principal-component structure under moderate missingness^22^. Complete-case sensitivity analyses reproduced the same loading pattern, confirming that the imputation did not bias the PCA solution. Sampling adequacy was confirmed (Kaiser–Meyer–Olkin = 0.89) and Bartlett’s test indicated sufficient shared variance (χ² = 783.3, p < 0.0001). Component retention followed Horn’s parallel analysis^23^: 5,000 Monte-Carlo permutations were generated, and components whose observed eigenvalues exceeded the 95ᵗʰ-percentile of the random distribution were retained. Scree-plot inspection (“elbow”) and Kaiser’s λ > 1 heuristic served as secondary, non-binding checks. No rotation was applied, preserving PC1 orthogonality for subsequent lesion-network mapping. Detailed loading vectors and explained-variance profiles are provided in **Supplementary Figure S3**; parallel-analysis curves are shown in **Supplementary Figure S4**.

### Lesion-Network Mapping (LNM) of Gait, Mobility, and Falls

LNM was conducted using the Lead Mapper from Lead-DBS (lead-dbs.org)^24^, as previously described. Briefly, for each patient, a functional connectivity profile was generated from their lesion mask using a normative resting-state fMRI connectome derived from 1,000 healthy individuals^25,26^. The resulting maps contain Pearson correlation coefficients, which were subsequently Fisher z-transformed.

First, separate LNM analyses were performed for the two behavioral clusters of interest: gait and mobility. For each, the PC1 from the respective domain was used as the independent variable. Statistical testing was performed using FSL’s randomise tool with 5,000 permutations and two contrasts: +1 for regions where connectivity was associated with impairment, and –1 for regions associated with spared function. Multiple comparisons were corrected using family-wise error (FWE) correction and threshold-free cluster enhancement (TFCE). Resulting T-score maps were overlaid on structural standard atlases to identify connected anatomical regions implicated in gait and mobility dysfunction, as described in detail previously^27,28^.

According to the description in a previous study^29^, spatial similarity scores for gait and mobility were derived from the uncorrected T-value LNM maps based on the individual connectivity fingerprints of each patient. Specifically, the voxelwise Pearson correlation coefficient between each patient’s lesion connectivity map and the symptom-specific network map was computed and then Fisher-z transformed. This quantified how strongly the spatial pattern of a patient’s lesion-related connectivity aligned with the respective symptom network. Higher spatial similarity scores, therefore, indicate a greater degree of overlap between an individual’s lesion connectivity pattern and the network associated with gait or mobility outcomes. Finally, to identify independent predictors of fall risk, we performed separate binary logistic regression analyses including age, treatment group (physical fitness vs. relaxation^14^), and either the mobility-related or gait-related spatial similarity score.

Lastly, we performed LNM for our primary outcome of interest: falls. Again, in this study falls refer to events reported during the intervention and follow-up period; importantly, no falls occurred during supervised intervention sessions. Fall ascertainment outside scheduled study visits was not systematic and relied on patient report at follow-up assessments. Fall occurrence was modeled as a binary dependent variable in FSL’s randomise tool, applying 5,000 permutations and two contrasts: [1, -1] to identify regions functionally connected to lesions associated with the occurrence of fall, and [-1,1] to identify regions connected to lesions in patients without falling. This analysis was adjusted for age, treatment group (based on the results of the prior binary logistic regression analysis), and baseline lesion volume. To evaluate the spatial similarity between the resulting fall network and the previously derived PC1-mobility and PC1-gait networks, we used voxel-wise Spearman rank correlation. Statistical significance was assessed with Moran’s spectral randomization (MSR) as implemented in the BrainSpace toolbox^30,31^. MSR exploits the eigenvectors of a spatial weight matrix to compare the observed correlation against a null distribution of spatially autocorrelated surrogate maps. We defined the spatial weights as the inverse squared inter-voxel distance to avoid arbitrary distance thresholds and to prevent over-weighting distant voxels. For each comparison, 10,000 surrogate maps were generated, and p-values were computed as the proportion of null correlations greater than or equal to the observed correlation^32–34^. All networks were down-sampled to 4-mm isotropic resolution for this analysis only, to improve computational efficiency while preserving spatial structure.

## Results

### Patient cohort description

A total of 110 patients were enrolled in the BAPTISe study^35^, of which 94 met our inclusion criteria. The mean age was 69 years (standard deviation [SD]: ±11), and the median NIHSS score at admission was 9 (interquartile range [IQR]: 6–12). A detailed summary of patient demographics, including cerebrovascular risk factors, stroke etiology, white matter hyperintensities, and lesion volume, is provided in **Supplementary Table S2**. A description of the characteristics for fallers (n = 17) is provided in **Supplementary Table S3.** A lesion heatmap illustrating the distribution of all ischemic brain lesions included in the analysis is shown in **Figure 2**. During the six-month follow-up period, 17 patients (18%) reported a fall. Fallers were older than non-fallers (mean age in years 75 vs. 67; p<0.05). Fallers had statistically significantly higher ARWMC scores compared to non-fallers (median 8,IQR 6-14 vs. 5, IQR 4-8; p<0.05). Lesion volume did not differ between fallers and non-fallers. Patient-reported mobility deficits were statistically significantly more common among non-fallers than fallers (88% vs. 92%, p=0.007).

**Figure 2.**
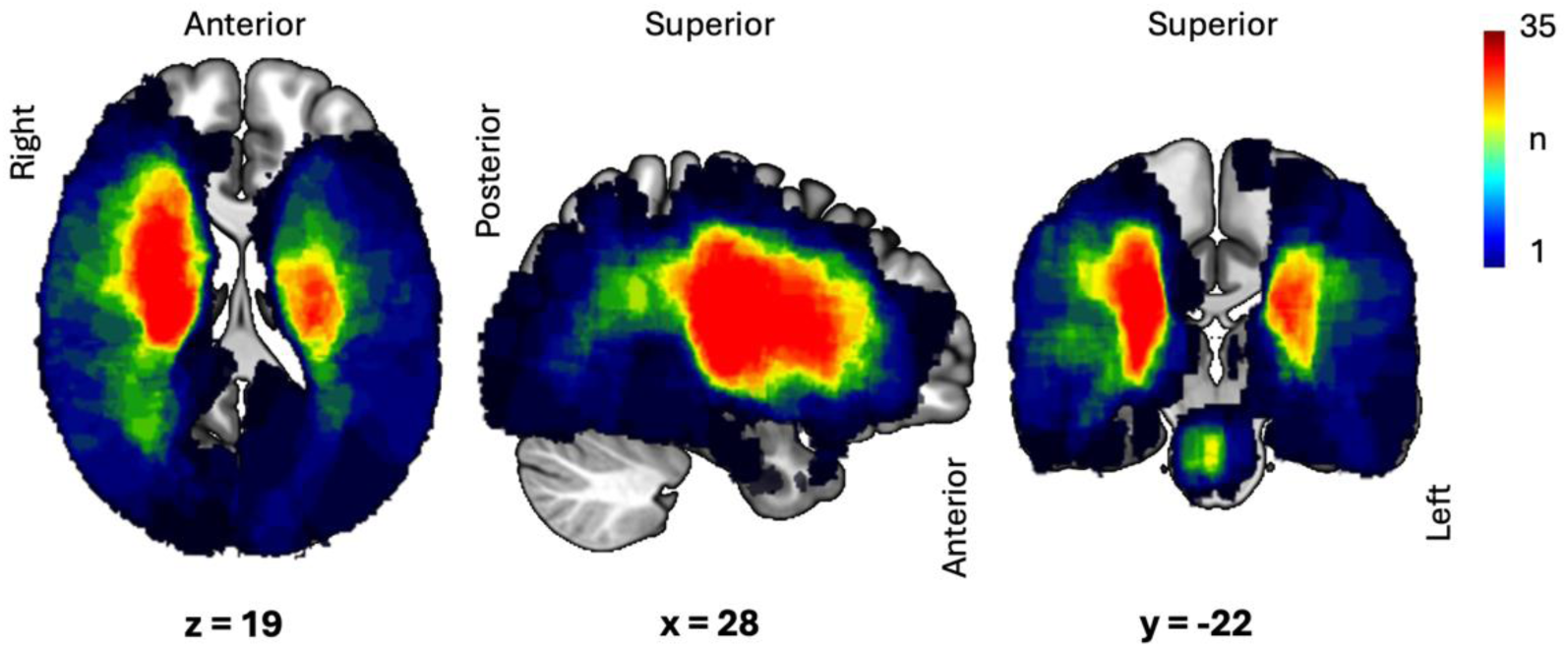
Heat-map showing lesion overlap from all patients included in the final analysis (N = 94) overlaid on a 100µm brain scan in MNI space. The color bar reflects the number of ischemic brain lesions overlapping at the voxel level, from blue to red, showing a maximal overlap of 35 lesions.

### Dimensionality Reduction of Gait and Mobility via PCA

In the mobility domain, PC1 accounted for 56% of the total variance; in the gait domain, PC1 explained 82.1%. All variables loaded positively on their respective PC1 (range: 0.31–0.53 for mobility, 0.47–0.53 for gait), indicating that higher scores consistently reflected greater functional impairment (**Supplementary Figure S3**). As anticipated, scatter plots confirmed strong positive correlations between each individual variable with the behavioral domains (mobility and gait) and the respective PC1 (all r ≥ 0.61, p < 0.001; **Supplementary Figure S1 and S2**). Horn’s parallel analysis (**Supplementary Figure S4**) confirmed that only the PC1 in both the mobility and gait domains exceeded the 95% confidence interval of the null distribution, supporting the retention of a single component in each domain for subsequent analyses. Visual inspection of scree plots revealed a clear elbow after the first component, and Kaiser’s criterion likewise supported the one-component solution.

### Mobility and Gait Networks from LNM and Fall Risk

Uncorrected T-score lesion network maps for both behavioral domains, mobility and gait, are shown in **Figure 3**. The PC1-Mobility network comprises predominantly cortical regions, including key sensorimotor areas such as the pre- and postcentral gyri, as well as associative and integrative regions like the superior and middle frontal gyri, insular cortex and supramarginal gyrus; 1-pFWE values ranged from 0 to 0.82. Notably, the PC1-Mobility network did not involve subcortical or cerebellar structures. The PC1-Gait network showed widespread involvement of subcortical regions – including the bilateral thalamus– as well as frontal cortical regions, including the precentral gyrus, paracingulate and cingulate cortices and insular cortex; 1-pFWE values ranged from 0 to 0.98. The network also included connectivity to infratentorial regions, including the brainstem and left cerebellar regions (left Crus I and II). For a full list of involved regions within the PC1-Mobility and PC1-Gait associated network, refer to **Table 1**.

**Figure 3.**
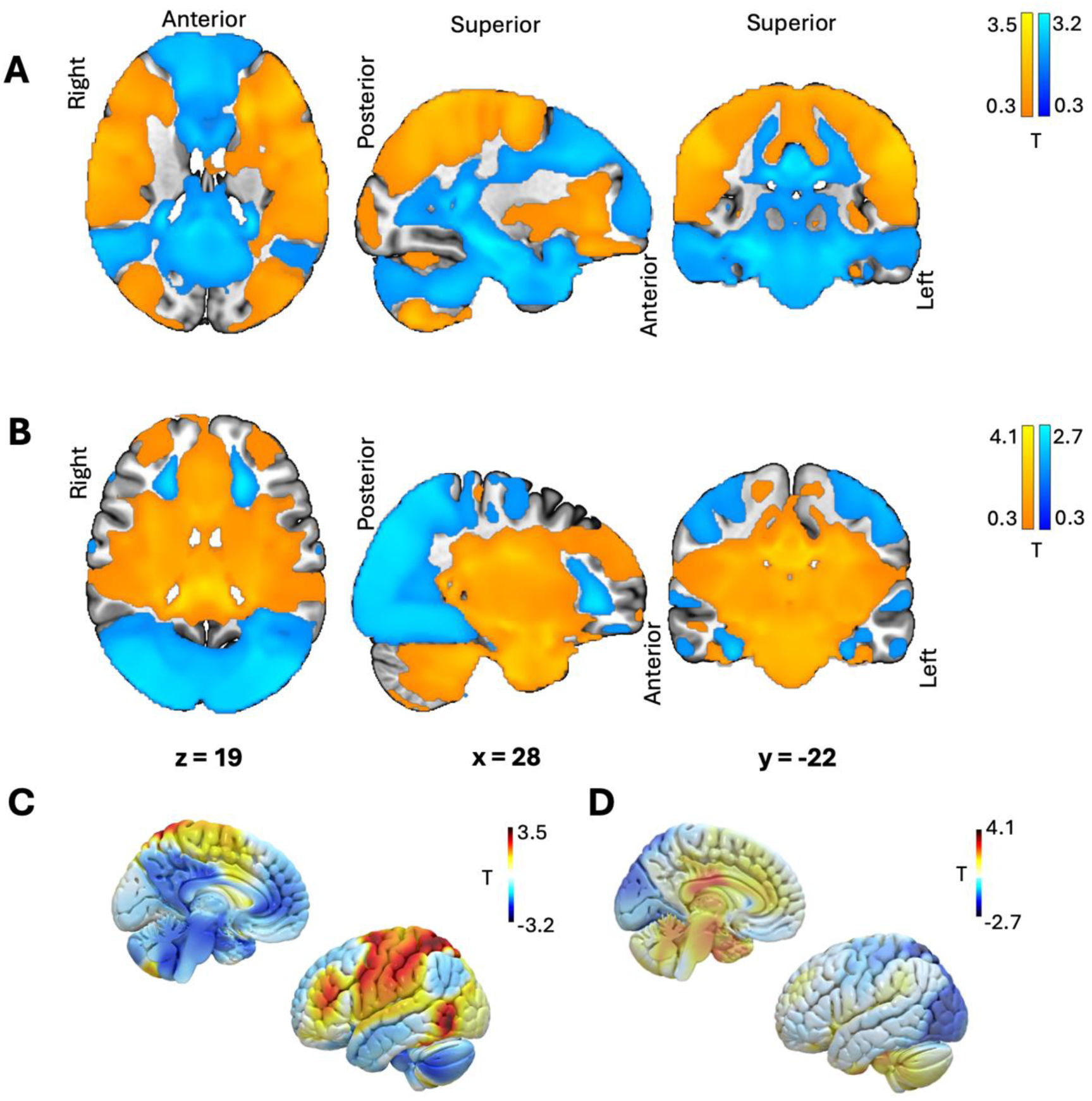
**Lesion-network maps for PC1-Mobility and PC1-Gait (N = 94). *(A)*** Uncorrected T-score map showing regions functionally connected to ischemic brain lesion locations associated with mobility impairment, using PC1 scores from the mobility domain as the continuous variable; 1-pFWE 0-0.82. Of note, these maps represent uncorrected T-score distributions and are presented for exploratory visualization purposes only; they do not reflect findings that survived FWE correction. ***(B)*** Uncorrected T-score map for gait impairment, based on PC1 scores from the gait domain (1-pFWE 0-0.98). Color bars indicate T-values, with red–yellow representing positive T values and blue–black representing negative T values. ***(C)*** Surface-rendered map of the mobility network (PC1-Mobility). ***(D)*** Surface-rendered map of the gait network (PC1-Gait). All maps are overlaid on the MNI152 100 µm brain template. Color bars indicate T-values, with values increasing from dark blue to dark red.

**Table 1.** Anatomical regions functionally connected to uncorrected T-score maps of PC1-Mobility and PC1-Gait. This table lists cortical, subcortical, and cerebellar regions identified within the functional symptom networks for PC1-Mobility and PC1-Gait. Regions were extracted using the Harvard-Oxford Cortical (HOOC) and Subcortical (HOOS) Structural Atlases, as well as the FNIRT-normalized Cerebellar Atlas, all in MNI152 standard space.

|  | HOOC | HOOS | Atlas of the Human Cerebellum |
| --- | --- | --- | --- |
| **PC1-Mobility Network** | Frontal Pole | -- | -- |
|  | Insular Cortex |  |  |
|  | Superior Frontal Gyrus |  |  |
|  | Middle Frontal Gyrus |  |  |
|  | Precentral Gyrus |  |  |
|  | Middle Temporal Gyrus, temporooccipital part |  |  |
|  | Inferior Temporal Gyrus, temporooccipital part |  |  |
|  | Postcentral Gyrus |  |  |
|  | Superior Parietal Lobule |  |  |
|  | Supramarginal Gyrus, anterior division |  |  |
|  | Supramarginal Gyrus, posterior division |  |  |
|  | Lateral Occipital Cortex, superior division |  |  |
|  | Lateral Occipital Cortex, inferior division |  |  |
|  | Juxtapositional Lobule Cortex (formerly Supplementary Motor Cortex) |  |  |
|  | Central Opercular Cortex |  |  |
|  | Occipital Pole |  |  |
| **PC1-Gait Network** | Frontal Pole | Left Thalamus | Left Crus I |
|  | Insular Cortex | Brain-Stem | Left Crus II |
|  | Superior Frontal Gyrus | Right Thalamus |  |
|  | Precentral Gyrus |  |  |
|  | Temporal Pole |  |  |
|  | Paracingulate Gyrus |  |  |
|  | Cingulate Gyrus, anterior division |  |  |
|  | Cingulate Gyrus, posterior division |  |  |
|  | Frontal Orbital Cortex |  |  |
|  | Central Opercular Cortex |  |  |

In univariable and multivariable binary logistic regression analysis for fall occurrence, advanced age (adjusted odds ratio [OR] 1.08, 95% Confidence Interval [CI] 1.02 – 1.15; p=0.01) was independently associated with increased falls in the models including either the PC1-Mobility or PC1-Gait spatial similarity score. Treatment group (i.e. intervention-arm physical activity vs. relaxation) was also associated with increased falls, as reported in the original trial results (adjusted OR 3.1, 95% CI 0.92 – 10.31, p=0.068), although statistically non-significant. Importantly, patient-specific mobility and gait spatial similarity scores were not associated with increased fall risk in either univariable or multivariable regression analyses (**Supplementary Table S4**). The multivariable model including the PC1-Mobility spatial similarity score showed slightly better overall fit (pseudo R² = 0.145, AIC = 83.67) compared with the model including the PC1-Gait spatial similarity score (pseudo R² = 0.139, AIC = 84.14), although the difference was negligible.

Although the PC1-mobility and PC1-gait network were not independently associated with fall risk in the regression analysis described above, we ran an additional, exploratory LNM analysis focused on the primary outcome of falls (N = 89) —adjusted for age, lesion volume, and treatment group. Here, fall occurrence after stroke was associated with lesions functionally connected to a bilateral network encompassing primary and secondary sensorimotor regions (pre- and postcentral gyri), parietal areas (superior parietal lobule and supramarginal gyrus), insular cortices, and the left cerebellar lobules, 1-pFWE values ranged from 0 to 0.21. Full atlas query results from the fall network are provided in **Supplementary Table S5**. Spatial similarity analysis using Moran’s I with 10,000 spatial surrogates demonstrated that the overlap between the uncorrected PC1-Mobility T-score map and the fall-associated network was greater than expected by chance (p < 0.001 and p = 0.477), as shown in **Figure 4**. In contrast, the spatial similarity between the uncorrected PC1-Gait and PC1-Mobility T-score maps, as well as between the PC1-Gait and fall-associated uncorrected T-score maps, was not statistically significant (both p > 0.05), as shown in **Supplementary Figure S5**.

**Figure 4.**
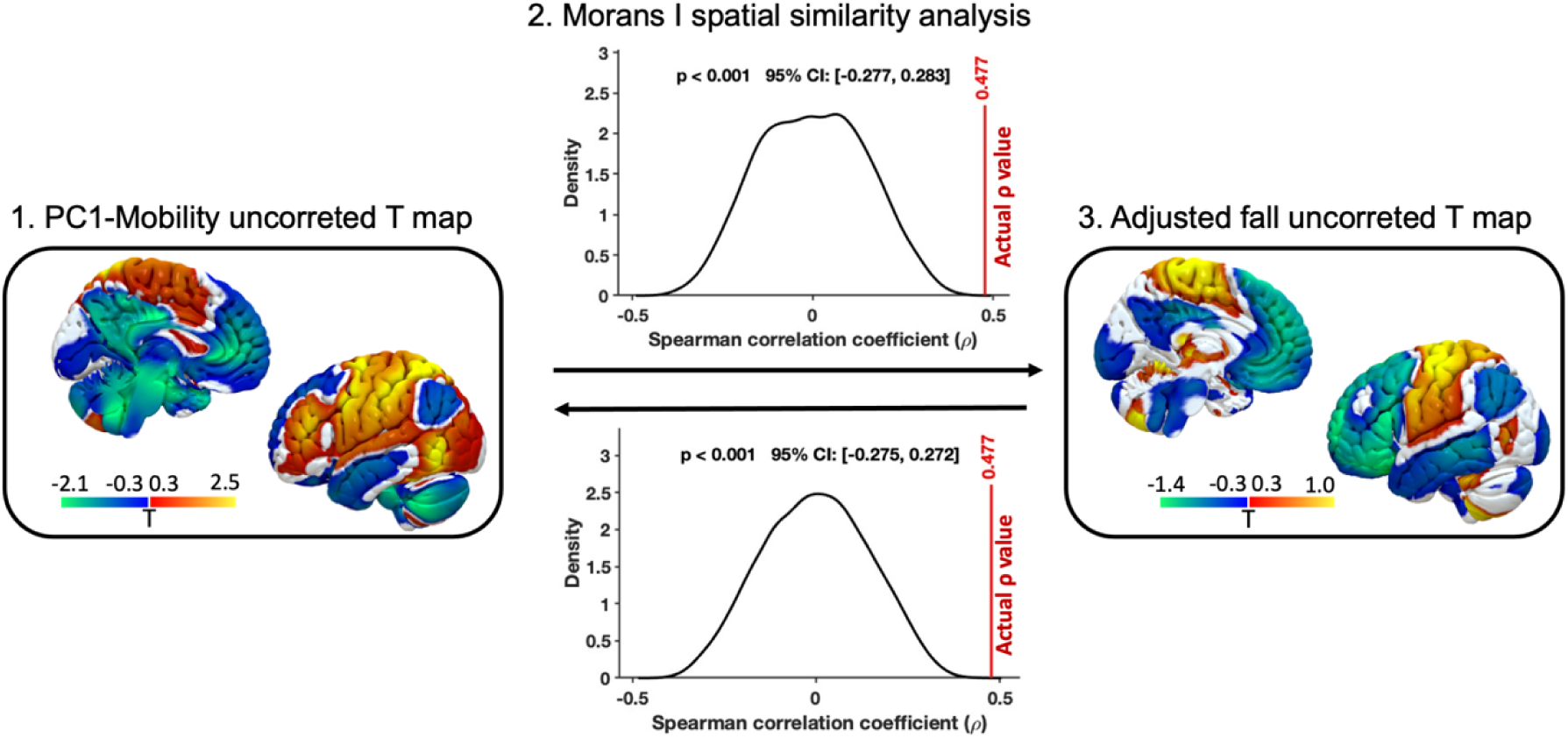
Similarity between PC1-Mobility brain network (N = 94) and adjusted fall network (N = 89). Whole-brain network similarity analysis using Moran’s I and 10,000 spatial surrogates to compare the spatial alignment between the network associated with PC1-Mobility and the network associated with fall adjusted for age, lesion volume, and treatment group. The map at the arrow origin is permuted, compared to the unpermuted target map. Network maps are statistically significantly more alike than expected by chance, indicating a high similarity. All maps are overlaid on the MNI152 100 µm brain template. Color bars indicate uncorrected T-values, with red–yellow representing positive connectivity and blue–green representing negative connectivity.

## Discussion

In this prospective cohort of patients with disabling ischemic stroke, we applied LNM to investigate the functional brain networks underlying mobility, gait, and falls, and their relationship to post-stroke fall risk. Impaired mobility was associated with lesions functionally connected to predominantly cortical regions, including primary and secondary sensorimotor areas, suggesting disruption of cortical motor execution and integration networks. In contrast, gait dysfunction was linked to lesions connected with subcortical and infratentorial structures—most notably the thalamus, brainstem, and cerebellum—highlighting the importance of subcortical circuits and cerebellar modulation in the coordination and automation of gait.

This study population consisted of patients with disabling ischemic stroke with moderate to high stroke severity (median NIHSS score of 9) and relatively large ischemic brain lesion volumes (median 11mL). Baseline clinical and demographic characteristics of the analyzed cohort (N = 94) were consistent with those reported in the main PHYS-STROKE trial^36^. Despite this, the overall fall occurrence was relatively low (18% within six months), compared to previous reports ranging from 30% to 50% up to one year^1,2,5^. This discrepancy is likely due to the short observation period but may also be attributed to the structured inpatient rehabilitation programs of patients within the BAPTISe cohort, which likely reduced fall risk through close clinical monitoring. Interestingly, patient-reported mobility deficits were statistically significantly more common among non-fallers than fallers (88% vs. 92%, p=0.007). This counterintuitive finding suggests that subjective awareness of mobility limitations may contribute to more cautious behavior, potentially reducing fall risk. Alternatively, it is possible that some non-fallers were more functionally impaired and therefore less ambulatory—e.g., reliant on wheelchairs or assisted transfers—reducing their overall exposure to fall risk.

Notably, 11 of the 17 fallers were either wheelchair-dependent or not mobile at all, compared to 57 of 77 non-fallers (**Supplementary T3**). Similarly, only one faller was able to transfer independently between bed and chair, versus 11 non-fallers. These findings suggest that limited mobility alone does not necessarily protect against falls. Interestingly, fallers also showed higher median spasticity scores (REPAS score: 12 [IQR 2–14]) compared to non-fallers (6 [IQR 2–11]), though this difference was not statistically significant (p = 0.396). This may indicate a trend toward greater motor impairment in fallers, but further studies are needed to clarify this association.

Fallers showed a higher white matter lesion load, as measured by the ARWMC score on baseline MRI (**Supplementary Table S2**). This association may be partially explained by the co-linear relationship between advanced age and increased white matter burden^37,38^. However, white matter lesions also reflect underlying small vessel disease and may disrupt critical motor and sensorimotor pathways—such as the corticospinal tract and thalamocortical loops—potentially contributing to impaired mobility and balance control after stroke^39,40^. Although the impact of white matter lesions on fall risk was not a primary aim of the current study, this association warrants further investigation in future studies.

While lesions to the corticospinal tract often result in hemiparesis or hemiplegia with characteristic hemiplegic gait patterns, damage to subcortical structures involved in motor modulation rather than direct motor execution, may preferentially impair gait control while leaving general mobility relatively intact.^8,41,42^ In line with prior work, mobility and gait were treated as related but distinct constructs in this study. Mobility refers to the ability to move freely and efficiently in everyday functional tasks such as transfers and short-distance ambulation, whereas gait encompasses walking speed, endurance, and coordination assessed under standardized conditions^43^. This distinction likely underlies the distinct symptom networks observed for the composite mobility score (PC1-Mobility) and the gait score (PC1-Gait; **Figure 3**).

In a binary logistic regression analysis for falls up to six months post-stroke, only advanced age was independently associated with increased falls (adjusted OR 1.08, 95% CI 1.02 – 1.15), which stands in line with previous studies^3,44^. As reported in the primary trial results, patients allocated to the physical fitness intervention group had a higher risk of falling compared to those in the relaxation group (adjusted OR 3.98, 95% CI 1.09-14.57 in PC1-Mobility model and adjusted OR 3.67, 95% CI 1.09-14.57 in PC1-Gait model). Similar rehabilitation trials in small stroke cohorts have also noted an increased frequency of adverse events, including falls, in intervention arms—highlighting the need for further investigation into the safety of early post-stroke mobilization strategies^45,46^. While we hypothesized that individual connectivity fingerprints to mobility and gait networks would increase fall risk, patient-specific spatial similarity scores were not independently associated with falls in this analysis (**Supplementary Table S4).** Given that falls were assessed during a defined rehabilitation phase in the current study and relied on patient self-report, caution is warranted in generalizing these findings to long-term post-stroke fall risk.

An additional network mapping analysis of fall occurrence identified a predominantly bilateral cortical network involving primary and secondary sensorimotor, parietal, opercular, insular, and occipital regions, with additional contributions from the right thalamus and left cerebellar lobule VI. Whole-brain spatial similarity analysis demonstrated a statistically significant overlap between this fall-associated network and the PC1-Mobility network (**Figure 4**). These findings are compatible with the hypothesis that post-stroke falls may be more closely linked to disruptions in cortical networks supporting voluntary motor control and sensorimotor integration than to subcortical systems primarily involved in automated gait modulation^41^.

This network-level finding aligns with clinical and experimental evidence indicating that post-stroke imbalance frequently arises from impaired multisensory integration, particularly of visual and somatosensory inputs^47^. Individuals with hemiplegia often rely more heavily on visual feedback to maintain postural stability, reflecting compensatory strategies that may be insufficient under complex or dynamic conditions. In parallel, hemiparesis-related gait abnormalities—such as reduced paretic propulsion, impaired single-limb support, and spatiotemporal asymmetries—necessitate energetically inefficient compensations that further increase fall susceptibility^12^. Emerging concepts of the somato-cognitive organization of the motor cortex provide an additional framework through which cortical lesions may disrupt higher-order motor planning and coordination, thereby contributing to fall risk beyond simple deficits in locomotor execution^41^.

Together, these observations support the notion that post-stroke falls are driven not solely by gait impairment, but by broader disturbances in cortical sensorimotor networks underlying balance, coordination, and whole-body control. This distinction may facilitate more refined fall-risk stratification and inform targeted secondary prevention strategies that extend beyond gait training alone.

## Strengths and limitations

First and foremost, it is important to note that this is an exploratory analysis and was not pre-defined in the statistical analysis plan of the BAPTISe study. Furthermore, information on falls was limited to a defined rehabilitation interventional phase; data on falls were binary (yes/no), without details on number, severity, or consequences (e.g., hospitalization). Additionally, the precise timing of each fall was not recorded, as falls were assessed only at follow-up visits via retrospective self-report. Furthermore, an inherent methodological limitation of LNM is the reliance on normative connectomes derived from healthy young adults, which may not fully capture functional connectivity patterns in older, neurologically impaired populations such as stroke survivors.

However, the study also offers notable strengths. It draws on a well-characterized cohort from a randomized controlled trial, with detailed and standardized assessments of gait performance and mobility. The inclusion of moderately to severely affected patients increase the likelihood of detecting meaningful lesion effects. Unlike prior LNM studies that rely on a single behavioral covariate, this analysis used PCA to reduce dimensionality across multiple clinical variables while preserving relevant clinical information. To our knowledge, this is the first study to investigate the prognostic value of data-driven gait and mobility networks in ischemic stroke patients concerning future fall risk.

## Conclusion

Stroke-specific contributors to fall risk remain poorly understood. In this exploratory study, we identified lesion-derived functional networks whose disruption may be more strongly associated with post-stroke falls—particularly networks involved in voluntary motor execution, action integration, and whole-body coordination—than those affecting subcortical regions modulating automated gait. These findings are hypothesis-generating and require validation in larger, independent cohorts. If confirmed, they may support lesion-informed risk stratification and guide more targeted fall prevention strategies during the early recovery phase.

## Data availability statement

Data supporting the results of this study can be requested by contacting the corresponding author.

## Code availability statement

Open source softwares were used for the pre-processing and analysis of the data, including: Lead-DBS (https://github.com/netstim/leaddbs), LESYMAP package in R version 4.2.0 (https://github.com/dorianps/LESYMAP), FSL version 6.0.6.4 (https://fsl.fmrib.ox.ac.uk/fsl/fslwiki/) and ANTsPy (https://github.com/ANTsX/ANTsPy).

## Author contribution statement

AKu, YT and ME jointly conceived the study. AKu, YT and UT designed the data analysis. YT and UT performed data analysis. AKu, YT, UT, AG, UG, DK, BW, AHN, MR, and ME interpreted the results. AK and YT wrote the manuscript. UT, TR, AHN pre-processed the MRI data. UG, TR, BW, AK gave conceptual and analytical advice. AHN and TR gathered neuroimaging data and helped with image pre-processing. U.G. helped with statistical analyses. All authors discussed the results and critically revised the manuscript.

## Acknowledgements

We acknowledge the use of OpenAI’s ChatGPT-4 for assisting in writing and correcting the code used in this study. The tool was used to enhance code efficiency and troubleshoot errors during the analysis. All scripts and generated outputs were critically reviewed and validated by the authors to ensure their accuracy and suitability for the research objectives.

